# Development of wastewater pooled surveillance of SARS-CoV-2 from congregate living settings

**DOI:** 10.1101/2020.10.10.20210484

**Authors:** Lisa M. Colosi, Katie E. Barry, Shireen M. Kotay, Michael D. Porter, Melinda D. Poulter, Cameron Ratliff, William Simmons, Limor I. Steinberg, D. Derek Wilson, Rena Morse, Paul Zmick, Amy J. Mathers

## Abstract

Wastewater-based monitoring for SARS-CoV-2 holds promise as tool to inform public health-decision making. Testing at individual building-level could be an efficient, passive means of preventing early detection of new cases in congregate living settings, but this approach has not been validated. Sample collection protocols were developed and refined during preliminary sampling from a hospital and a local municipal wastewater treatment plant. Molecular diagnostic methods were compared side-by-side to assess feasibility, performance and sensitivity. Optimized sample collection and processing protocols were then used to monitor two occupied dormitory complexes (n=105 and 66) over eight weeks. Wastewater results were validated using known case counts from external clinical testing of building occupants. Results confirm that ultracentrifugation from a 24 hour composite collection had a sensitivity of 95% and a specificity of 100%. However, if the detection of convalescent shedding is considered a false positive then the sensitivity would be 95.2% but the specificity would drop to 52%. We determined a highly sensitive method for detecting SARS-CoV-2 shedding in building wastewater however our methods could not distinguish new infectious cases from persistent convalescent shedding of SARS-CoV-2 RNA. Future work must focus on methods to distinguish new infections from convalescent shedding to widely deploy this promising wastewater surveillance tool.

## Introduction

Severe Acute Respiratory Syndrome Coronavirus 2 (SARS-CoV-2) is a highly contagious respiratory virus with a case-fatality rate of 1-2% ^1^. Mask-wearing has been shown to reduce the spread of SARS-CoV-2, thereby decreasing the incidence of Covid-19, the illness caused by SARS-Cov-2 ^2^. Because people are unlikely or unable to wear masks at all times while they are in their homes, congregate living settings (e.g. nursing homes, prisons, etc.) have been disproportionately impacted by large outbreaks in the pandemic thus far ^3-6^. Approaches to limit transmission in such settings have included aggressive entry screening via wellness attestations and/or temperature checks ^7, 8^; however, the effectiveness of these strategies has been hampered by the fact that an infected individual is contagious prior to or without ever exhibiting symptoms ^9-11^. To prevent outbreaks in congregate living settings, there is urgent need for frequent surveillance to quickly detect new cases and isolate them from other occupants ^12, 13^. Frequent testing, of all occupants could be an effective means of identifying and separating contagious individuals and quarantining others ^3, 4, 14^. However, there are significant logistical considerations associated with testing all occupants repeatedly; cost, personnel, test availability, privacy, compliance, testing fatigue, occupant anxiety^15^.

Stool has high concentrations of SARS-CoV-2 RNA early in infection with persistent shedding at lower concentrations for weeks after infection ^16, 17^. Some groups have been able to recover live virus from stool, but feces appears to contain predominantly non-viable RNA fragments, at surprisingly high concentrations ^16, 17^. Fecal detection of SARS-CoV-2 RNA, coupled with the strong need for passive routine surveillance, has prompted strong interest in wastewater-based testing (WBT) as means to monitor for prevalence of the virus ^16, 18^. Recent published work documents the potential usefulness of this strategy at a community level (e.g., via sampling at a municipal wastewater treatment facility) ^19-24^. A working group comprising water and wastewater professionals from academia and public practice have asserted that, “wastewater surveillance in sewersheds is a rapidly developing area of research that has the potential to inform public health policy decisions in the context of the current pandemic.” ^25^.

There is rapidly emerging interest in wastewater-based monitoring for SARS-CoV-2 RNA at the individual building level for various congregate living settings (e.g., prisons, university dorms) ^26, 27^. It is anticipated that pooled wastewater-based testing of could act as early surveillance of all building occupants. Wastewater results could be used to help determine where and when point prevalence and/or other public health interventions are warranted, as means to prevent further spread of Covid-19 but avoid the logistic barriers of frequent massive individual testing. There have been popular (lay) press reports about how this approach has been implemented for various university dormitories; e.g., at the University of Arizona, Syracuse University, and Pennsylvania State University. However, there is no clarity or standardization among the protocols that have been used thus far, nor is there any agreement on what constitutes best practices for implementation of this approach ^28^. Among this group, work from Syracuse University offers the greatest amount of technical detail, but additional investigation and external validation are still needed ^27^.

The overarching objective of this work was to evaluate the hypothesis that frequent monitoring of pooled wastewater samples could be an efficient means of monitoring for Covid-19 cases among building occupants. Three specific objectives were pursued: 1) establishing a robust strategy for collecting representative wastewater samples from occupied congregate living settings; 2) comparing and optimizing molecular diagnostic techniques; and 3) validating results based on external data. We collected wastewater samples from several populations of individuals undergoing frequent clinical testing for Covid-19; in a hospital and several occupied university dormitories. We also analyzed municipal water and wastewater samples.

## Methods and Materials

### Sampled Settings

#### Hospital

The hospital setting was a newly constructed tower at University of Virginia (UVA) Medical Center. This tower contains 84 rooms over three occupied floors but was only at roughly 40% occupancy during this study. The new tower opened in early April 2020 and during the study period it was used for isolation and/or treatment of only Covid-19 patients or individuals needing to quarantine after exposure to Covid-19. Both acute care and intensive care unit Covid-19 patients are housed in this location. All rooms are single occupancy, with separate attached-bathroom harboring a toilet, a sink and enclosed by a door. Mobile patients use the toilets. Based on infection control guidance, and in consultation with nursing staff, the protocol is to dispose of stools from immobile patients (via rectal tube, bedpan, or commode) via the toilet within each room. However, small stools are sometimes captured onto pads that are disposed of into the solid waste stream. Dye-based point source tracking was performed to confirm that candidate wastewater sampling locations were hydraulically connected with the desired patient rooms as well as measure transit time. Approximately 100 mL of a commercial liquid dye (FLT/Yellow-Green, Bright Dyes, Miamisburg, OH) was introduced via a patient-room toilet or solid waste hopper. The toilet was then flushed continuously for 2.5 min to push the dye through the wastewater collection system. Visual monitoring at downstream locations was used to confirm flow connectivity and estimate transit times.

#### Occupied dormitories

The dorm settings were two complexes located approximately 0.5 km apart from each other. Both complexes contain multiple apartment-style buildings. Complex A comprises five individual buildings, each containing 20 units. Each unit contains two single-occupancy bedrooms, one and a half bathrooms, and shared kitchen. Complex B comprises three individual buildings, each containing 15 units. Each unit contains four single-occupancy bedrooms, a single shared bathroom, and a single shared kitchen. Neither location was at full occupancy during the study interval. Many of the building occupants were student-athletes who were required to wear masks and encouraged to practice social distancing and good hand hygiene. Enhanced cleaning protocols were used in the dormitories to avoid widespread transmission.

#### Wastewater treatment plant (WWTP)

The Moores Creek Advanced Water Resource Recovery Facility in Charlottesville, Virginia is a municipal wastewater treatment plant (WWTP) operated by the Rivanna Water and Sewer Authority (RWSA). Its design flow rate is approximately 15 million gallons per day (MGD). Its service population is approximately 143,000 including UVA. It serves the City of Charlottesville plus some portions of the surrounding Albemarle County.

#### Private residence

Municipal tap water was collected from a private residence (four full time occupants) located within the same distribution network that serves the UVA Hospital and sampled dormitories.

### Sampling protocols

We adhered to CDC protocols pertaining to safe handling of sanitary wastewater during all sampling events, with the following modifications ^29, 30^. All operators wore cloth or paper non-N95 masks, protective eyewear or a face shield, a disposable liquid impermeable gown, and gloves. Alcohol based hand hygiene was practiced before and after sampling. All equipment surfaces that came into direct contact with the wastewater were flushed with a 10% bleach (vol/vol) solution. Contact times during equipment sanitization was at least 5 min, followed by multiple rinses with tap water to remove bleach residue. Care was taken to have the collection done principally by a single person wearing full PPE, assisted by another person. Earliest samples from the UVA hospital constituted grab samples, whereby a small container was attached to a long pole and held underneath the outfall to collect volumes of 5-6 liters, over a period of 10-15 minutes.

For composite samples autosamplers (Sigma 900 Max and AS950) (Hach, Loveland, CO) were employed with running a time-weighted sampling program over periods of approximately one day (20-24 h). Several different sampling programs were attempted, to balance battery life with sampling frequency over the desired interval. Time interval between successive samples was 10-30 min between successive samples, whereas collected volumes were 30-50 mL per sampling event. For the majority of the study, the standard program was 30-mL samples collected every 15 min. Under especially low flow conditions, it was sometimes necessary to tape over parts of the sample collection probe (strainer) to exclude air and improve sample draw. Auto-samplers were packed with ice during sample collection. At the end of each sampling interval, samples were immediately transported to a laboratory and processed the same day.

Rivanna Water and Sewer Authority personnel provided 1-L samples of their composited raw influent as well as 50-mL samples of the solids from the underdrain of their primary clarifiers (“primary solids”). Compositing starting at 12 am, and samples were collected at roughly 10 am. All samples were transported to lab on ice and processed on the same day that they were collected.

Approximately 60 mL of tap water from a private residence were collected 1-2 times per hour over a 24-hour period. The sample was refrigerated during collection, transported on ice to the laboratory, and processed the same day.

### Molecular Methods

Four wastewater RNA concentration methods and two extraction assays for analysis of SARS-CoV-2 in wastewater matrix were assessed. The four concentration methods were: 1) electropositive filtration ^31^, 2) ultracentrifugation with a sucrose gradient ^27^, 3) polyethylene glycol (PEG 8000) precipitation ^32^, and 4) an alternative PEG precipitation based on a protocol from the MIT Alm Lab and Biobot Analytics ^19^.

For the electropositive filtration method, 6 liter of wastewater were passes through a ViroCap™ filter (Scientific Methods, Granger, IN, USA) to collect negatively charged viral particles. The viral RNA was then eluted in a solution of 1.5% beef extract and 0.05 M glycine (pH = 9.5) and subjected to a secondary concentration step via flocculation in a 5% skim milk solution ^28^.

For the ultracentrifugation method, 40 mL of a mixed wastewater sample was added to an ultracentrifuge test tube (Beckman Coulter, Indianapolis, IN)). Then 24 mL of a concentrated sucrose solution (50% sucrose in TNE buffer) was carefully pipetted underneath the sample to create two distinct layers. Samples were spun down in a Beckman Coulter LE80 Ultracentrifuge for 45 minutes at 42,000 rpm (∼150,000 x g). Supernatant was decanted, and pellet was resuspended in 300 mL PBS^27^.

The third and fourth concentration methods utilized polyethylene glycol (PEG 8000) to precipitate virus particles. To apply the PEG precipitation method of Hjelmsø et al, 200 mL of mixed wastewater was combined with 25 mL of glycine buffer (3% beef extract, 0.05 M glycine, pH = 9.6). The mixture was then centrifuged at 8,000x g for 30 min, and the supernatant was filtered using a 0.45-μm PES membrane. PEG 8000 (80 g/L) and NaCl (17.5 g/L) was added to the filtered supernatant and incubated with the mixture overnight at 4 °C under agitation. Samples were the centrifuged at 13,000xg for 90 min. The resulting pellet was resuspended in 1 mL PBS. By the fourth method (Biobot) 120 mL mixed wastewater was filtered through a 0.22-uM filter and combining the filtrate with 12g PEG 8000 and 2.7g of NaCl. The sample was shaken for 15 min to fully dissolve PEG/NaCl and then centrifuged at 12,000g for 1 h. Supernantent was discarded and sample was re-centrifuged at 12,000g for 5 min to collect particulates. Resulting supernatant was decanted from the tube, and the pellet was re-suspended in Trizol.

The RNA concentrated pellets from all methods were then processed using two commercial RNA extraction kits: QiaAmp Viral RNA Mini Kit (Qiagen, Hilden, Germany), and NucleoSpin RNA Plus Kit (Takara Bio USA Inc, USA). Manufacturers’ protocols were followed as written, with the addition of a dithiothreitol (DTT) pretreatment step for the QiaAmp kit to attempt to decrease PCR inhibition. For the pretreatment step, DTT, 500mM final concentration, was added at equal volumes to sample volumes and vortexed thoroughly. Mixtures were then incubated at room temperature for 30 min with intermittent vortexing. For all experiments, the PCR assay was run on the ABI 7500 Fast Real Time PCR system (Applied Biosciences) using primers and methods to amplify RNAseP (Rp) (PCR human control), N1 and N2 SARS-CoV-2 viral targets, as specified in the CDC protocol for SARS-CoV-2 analysis ^33, 34^. We recorded cycle threshold (Ct) values for all analyses. All runs have a positive, negative and water blank control. The criterion for a positive result was the same the cutoff for clinical diagnostic testing; namely, that Ct values for N1, N2 were ≤45. The RP control was present to evaluate for inhibition but if positive and negative control were successful a positive could be determined with a negative Rp. Negative samples corresponded to Ct values >45 for N1 and N2, with RP ≤ 45 and indeterminate were when only one target was positive.

### Results Validation and Interpretation

Most patients present in the Covid-19 tower on hospital sample days had undergone SARS-CoV-2 PCR testing prior to admission or when they were first admitted. This testing could have been performed by UVA’s in-house clinical laboratory (Cepheid Infinity, Abbott m2000 or Abbott Allinity), or at an outside hospital on another platform. Individual patient charts were reviewed to collect the following information: date of initial positive Covid-19 test and date of symptom onset and total tower occupancy (IRB# 22521).

For the sampled dormitories, all occupants were required to undergo midturbinate nasal swab SARS-CoV-2 PCR testing (Abbott Alinity or m2000 Chicago, Il) (weekly Complex A or biweekly Complex B) throughout the study interval. Once an occupant tested positive they were not re-tested. The individual then isolated for 14 days starting from the test result date. These tests were not undertaken as part of this study. The research team did not have access to individual charts for dorm occupants but were provided with testing results in aggregate form, thus this was determined to not be human subjects research.

Sensitivity analysis was done with number of known occupants in a building or catchment as the reference for test performance ^35^. We assumed that one or more persons in a building who were in the infectious period of 0-14 days from PCR positivity with or without symptoms would be the reference method for positive^18^. Basic statistical calculations were performed using the Microsoft Excel Data Analysis Toolpak (Excel 2019).

## Results

### Preliminary Sampling and Methods Comparison – Hospital and WWTP

After several repeated, definitively negative results from the hospital, mostly collected by grab sample, dye testing was used to confirm hydraulic connectivity between patient rooms and candidate testing sites. Two early dye tests in the hospital tower revealed that wastewater from the Covid-19 unit was not accessible via several manholes initially considered to be promising sampling locations. This outcome prompted a third set of dye tests in a new outflow location, in which it was observed that transit time from a toilet in an unoccupied patient room to several accessible candidate location was approximately 2-3 min. Travel time from a soiled waste hopper on the same floor was also in this time range. Grab samples collected from manholes downstream of the Covid-19 tower, where its wastewater commingles with flow from the rest of the hospital, yielded inconclusive results (data not shown). Therefore, all subsequent hospital samples were collected from a cleanout valve located just upstream of where the wastewater exits the building. Overnight composite samples (typically 22-24 h intervals) were collected from this location with 30 mL every 15 minutes as the typical setting. Average daily flow rate for days on which hospital samples were collected was 9.0-13.5 gallons per minute.

Composite samples of hospital wastewater, as well as raw influent and primary solids from the WWTP, were used to compare SARS-CoV-2 published concentration and extraction protocols ^23, 36-39^. Results are summarized in Table 1. An immediate observation from these results is that SARS-CoV-2 was detectable in wastewater from individual buildings and WWTP samples although the various methods had variable ability to detect.

**Table 1.** Ct values for preliminary samples from hospital cleanout valve and the local municipal WWTP, as processed using various concentration and extraction methods from literature. Smaller Ct values are potentially indicative of better SARS-CoV-2 recovery. All samples were collected on July 7, 2020 and processed the same day. Gray shading indicates “positive” determinations. Blanks correspond to combinations of protocols that were not assessed.

| Source | Extraction | C(t) vlaue |  |  |  |  |  |  |  |  |  |  |  |  |  |  |
| --- | --- | --- | --- | --- | --- | --- | --- | --- | --- | --- | --- | --- | --- | --- | --- | --- |
|  |  | EP Filtration |  |  | Ultra-centrifugation<sup>1</sup> |  |  | PEG Precipitation |  |  | BioBot PEG |  |  | No Concentration |  |  |
|  |  | N1 | N2 | RP | N1 | N2 | RP | N1 | N2 | RP | N1 | N2 | RP | N1 | N2 | RP |
| Hospital | None | 31.18 | 32.82 | 33.27 |  |  |  |  |  |  |  |  |  |  |  |  |
|  | QiaAmp | 33.39 | 33.24 | ND* | 33.25<br>36.46 | 39.85<br>34.71 | 35.31<br>32.62 | ND | 35.17 | ND | ND | ND | ND |  |  |  |
|  | NucleoSpin | 28.94 | 29.47 | 31.01 | 30.90 | 32.54 | 30.14 | 31.41 | 31.28 | ND* | 36.7 | 37.11 | 35.11 |  |  |  |
| WWTP Raw Influent | QiaAmp |  |  |  | ND<br>38.19 | ND<br>ND | 34.4<br>232.94 | 35.51 | ND | ND | ND | ND | ND |  |  |  |
|  | NucleoSpin |  |  |  | 30.26 | 31.23 | 28.32 | 33.69 | 32.54 | 33.27 | ND | ND | ND |  |  |  |
| WWTP Primary Solids | QiaAmp |  |  |  |  |  |  |  |  |  |  |  |  | 36.22 | ND | ND |
|  |  |  |  |  |  |  |  |  |  |  |  |  |  | 34.75 | 36.21 | 40.1 |
|  | NucleoSpin |  |  |  |  |  |  |  |  |  |  |  |  | 31.76 | 32.46 | 34.62 |

<sup>1</sup>Italicized font indicates post-extraction treatment with DTT prior to q-PCR. Not detected (ND) (Ct≥45) Unitalicized font for same processing conditions corresponds to identical sample without DTT treatment.\*Demonstrates sample inhibition but positive for the presence of SARS-CoV-2 RNA

Although only compared at a single time point, of the four evaluated concentration protocols, ultra-centrifugation demonstrated less inhibition and more positive results for both hospital wastewater and WWTP raw influent during preliminary sampling. The third wastewater, WWTP primary solids, did not require concentration because it rapidly self-settled by gravity, making it straightforward to collect a pellet. In contrast, each PEG protocol had lower yield with a positive result for both sample types sample types but only with the Nucleospin extraction and the Ct values were higher compared to the Ultracentrifuge method with the same extraction^32^, and WWTP raw influent for the BioBot technique^19^ did not work on the WWTP with either extraction method but worked on the hospital with the Nucelospin extraction but with higher Ct values. The electropositive filtration technique resulted in 2 positives and 1 inhibited sample across the tested hospital sample.

Regarding extraction, the NucleoSpin protocol yielded 7 positive results out of 8 trials during side-by-side methods comparison. The positives were spread across all three kinds of evaluated samples (hospital wastewater, WWTP raw influent, and WWTP primary solids). In contrast, the QiaAmp protocol with and without DTT yielded only 3 positive results out of 11 trials (Fischer exact p=0.02), and none of the positives corresponded to WWTP raw influent samples. Six out of the 11 QiaAmp and one of the 15 (Fischer exact p=0.02) extracted samples failed internal control QC, which monitors sample inhibition of the enzymatic PCR reaction. Post-extraction treatment of the QiaAmp samples with DTT improved SARS-CoV-2 detectability; i.e., decreasing Ct value compared to an identical sample without DTT, and transforming what would have been an inhibited/failed sample into a positive result. However, for samples treated using the same concentration protocol, it was observed that NucleoSpin extraction gave lower Ct values than QiaAmp extraction followed by DTT treatment.

Based on results from the side-by-side method comparison (Table 1) and other factors (see discussion), ultra-centrifugation followed by NucleoSpin extraction was selected for continued use in this study. Additional samples were then collected from the hospital and WWTP over a period of several weeks. Table 2 summarizes resulting Ct values for these samples, plus one sample of municipal tap water collected from a location within the distribution network servicing the UVA hospital and dorms. Table 2 also presents relevant Covid-19 case count comparators for each hospital and WWTP sampling date; namely, corresponding patient census data and known cases within the WWTP catchment area, respectively.

**Table 2.**
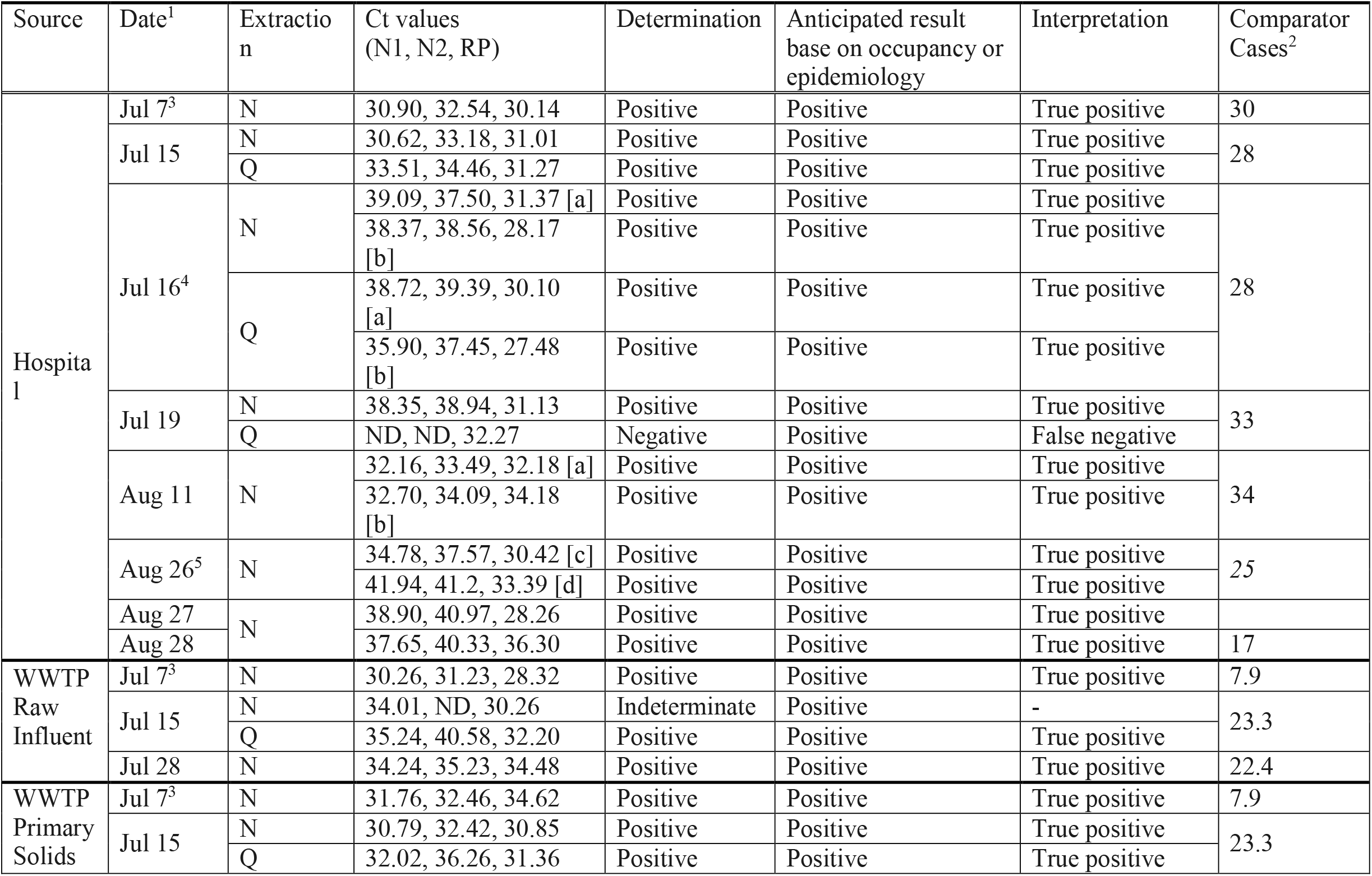
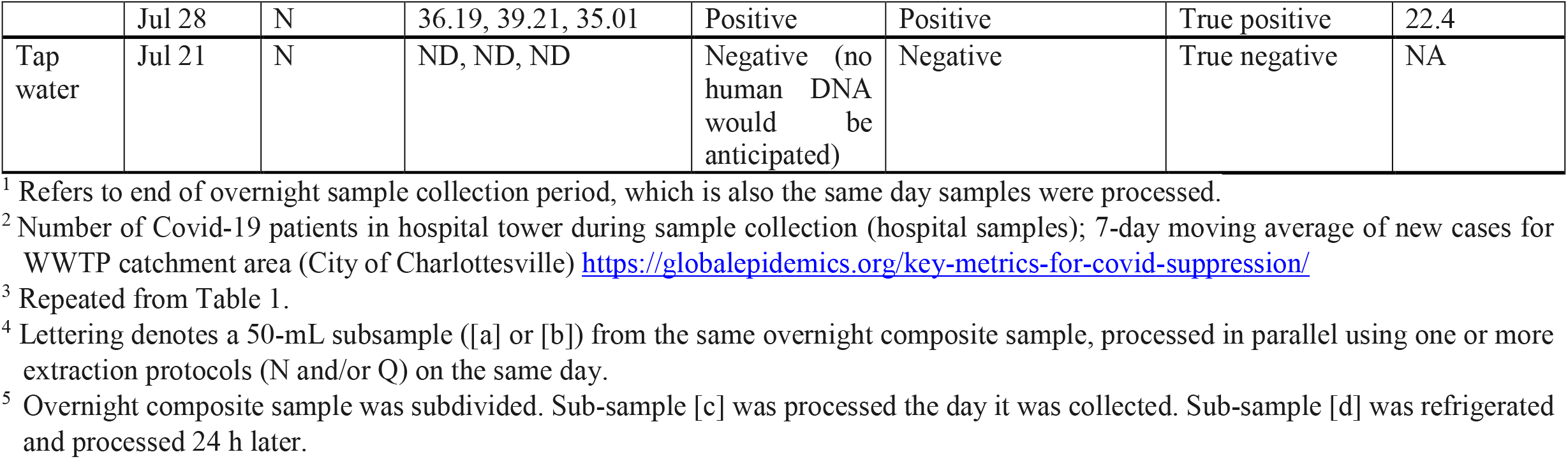
Sampling results for UVA hospital and the local municipal WWTP over time. Lower Ct values indicate higher detected concentrations of SARS-CoV-2. Extraction methods are NucleoSpin (N) and QiaAmp (Q). Not detected (ND), which indicates Ct >45.

### Sampling from Individual Occupied Dormitories

GIS plans were used to identify sampling locations within two occupied dorm complexes. Figure 1A shows the locations of three sampling sites within Complex A: one manhole collecting flow from all five buildings (red star), plus two building-level cleanout pipes (labeled “A” and “R” on Buildings 4 and 5, respectively). Figure 1B shows the location of the single sampling site in Complex B, which was a manhole collecting flow from all three buildings in the complex.

**Figure 1.**
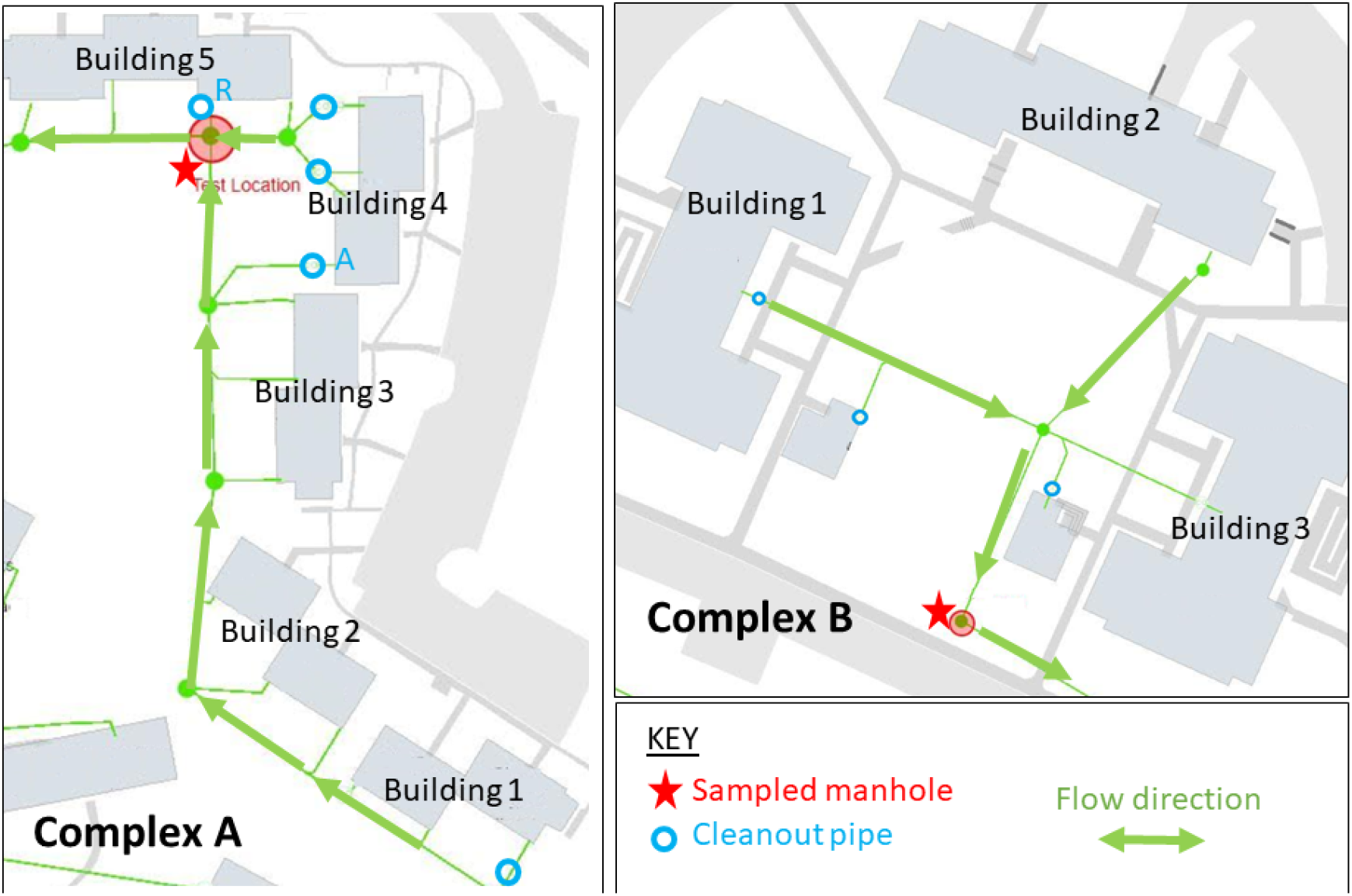
Maps of UVA dorms in Complex A (left) and Complex B (right). Red stars denote each sampled manhole. Arrows indicate flow directions. Cleanout valves labeled “A” and “R” denote secondary testing locations (via cleanout pipes) for selected buildings in Complex A.

Thirteen samples were collected from Complex A, corresponding to 0-3 samples per week over 8 weeks. Approximately 105 occupants were living in Complex A where the wastewater was collected as the manhole test collection during this period. Four samples were collected from Complex B, corresponding to 0-2 samples per week over four weeks. Sixty-six occupants were living in Complex B during this period and all wastewater was collected at the sampled manhole. Table 3 summarizes Ct values for samples from both complexes, alongside known case counts by week.

**Table 3.**
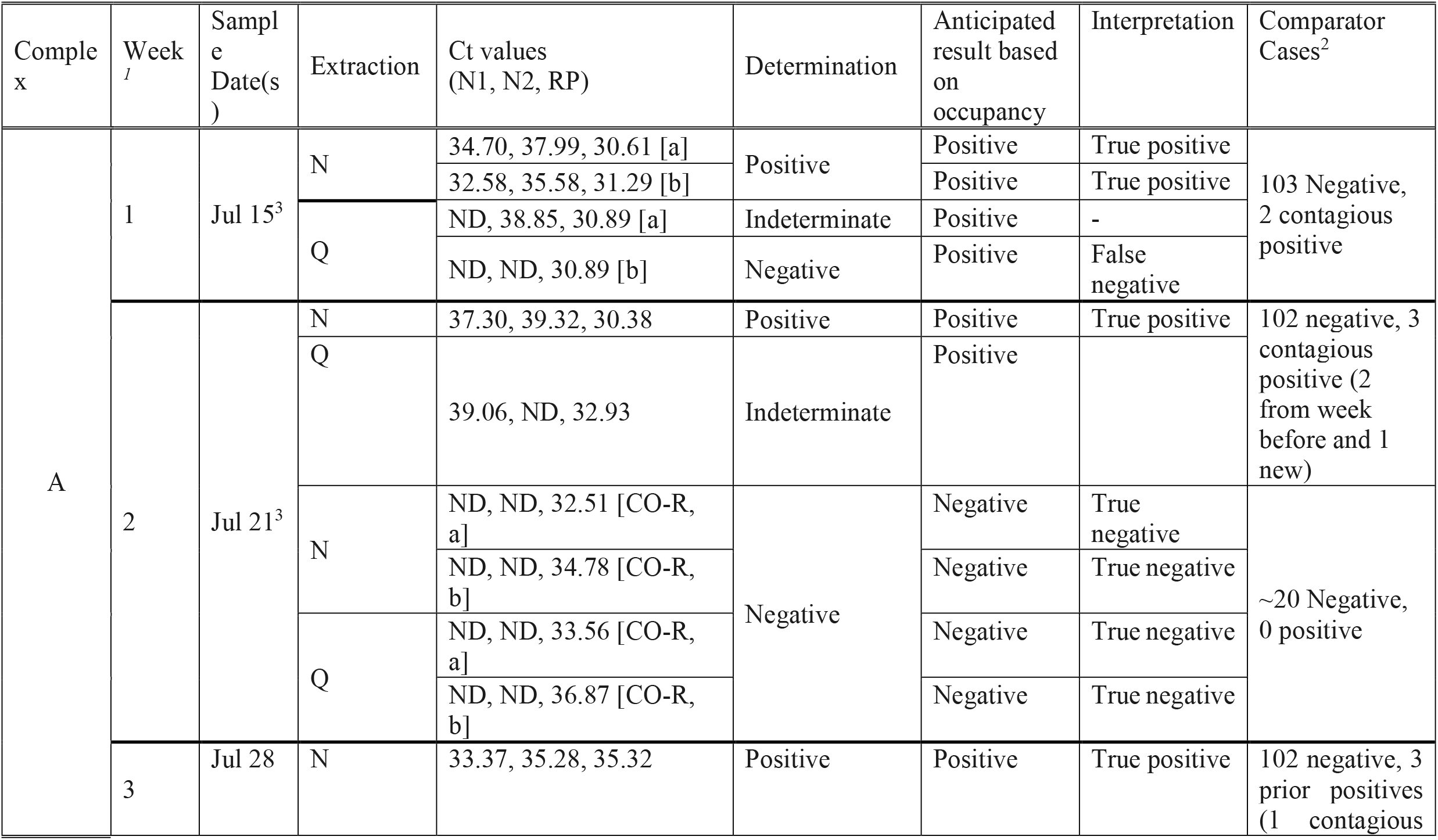
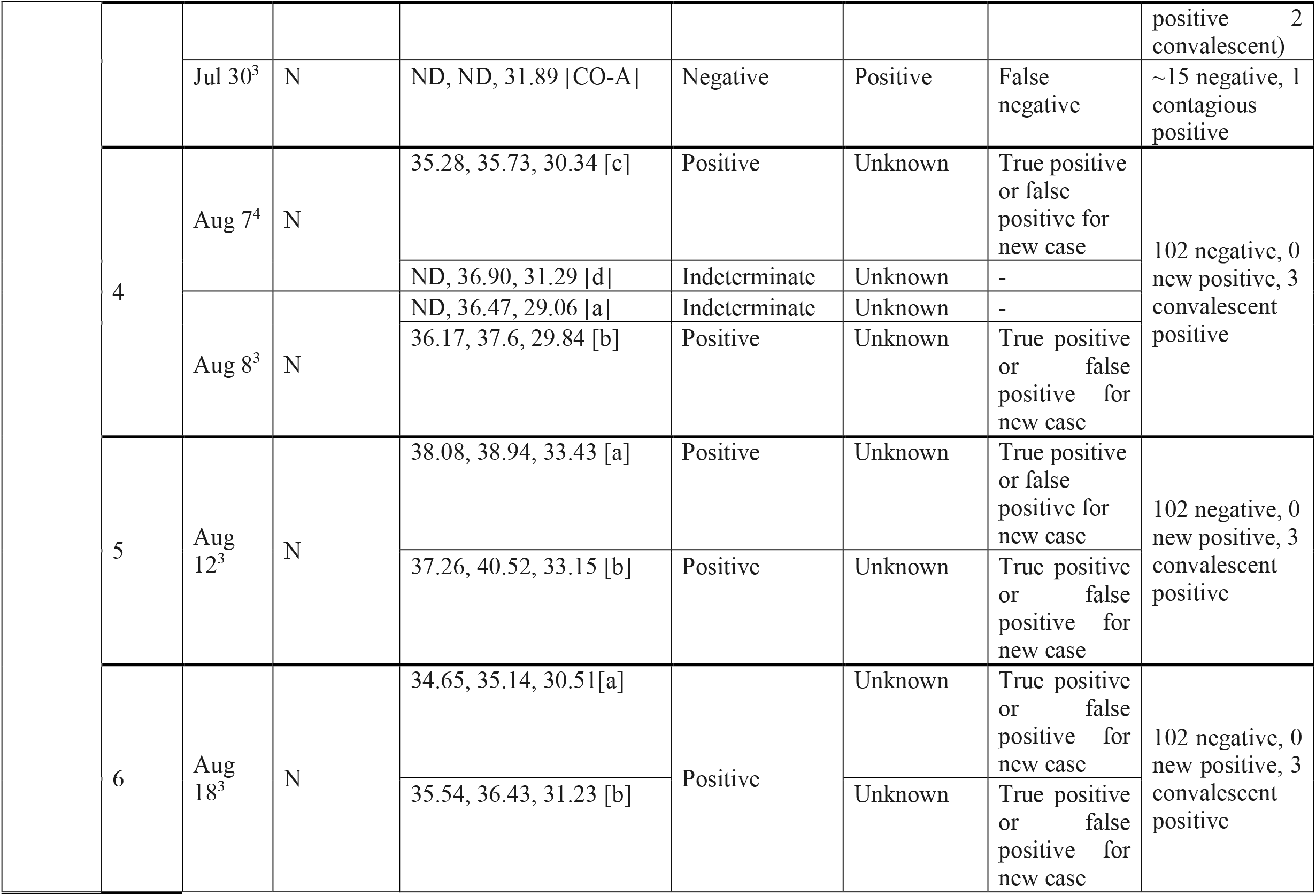
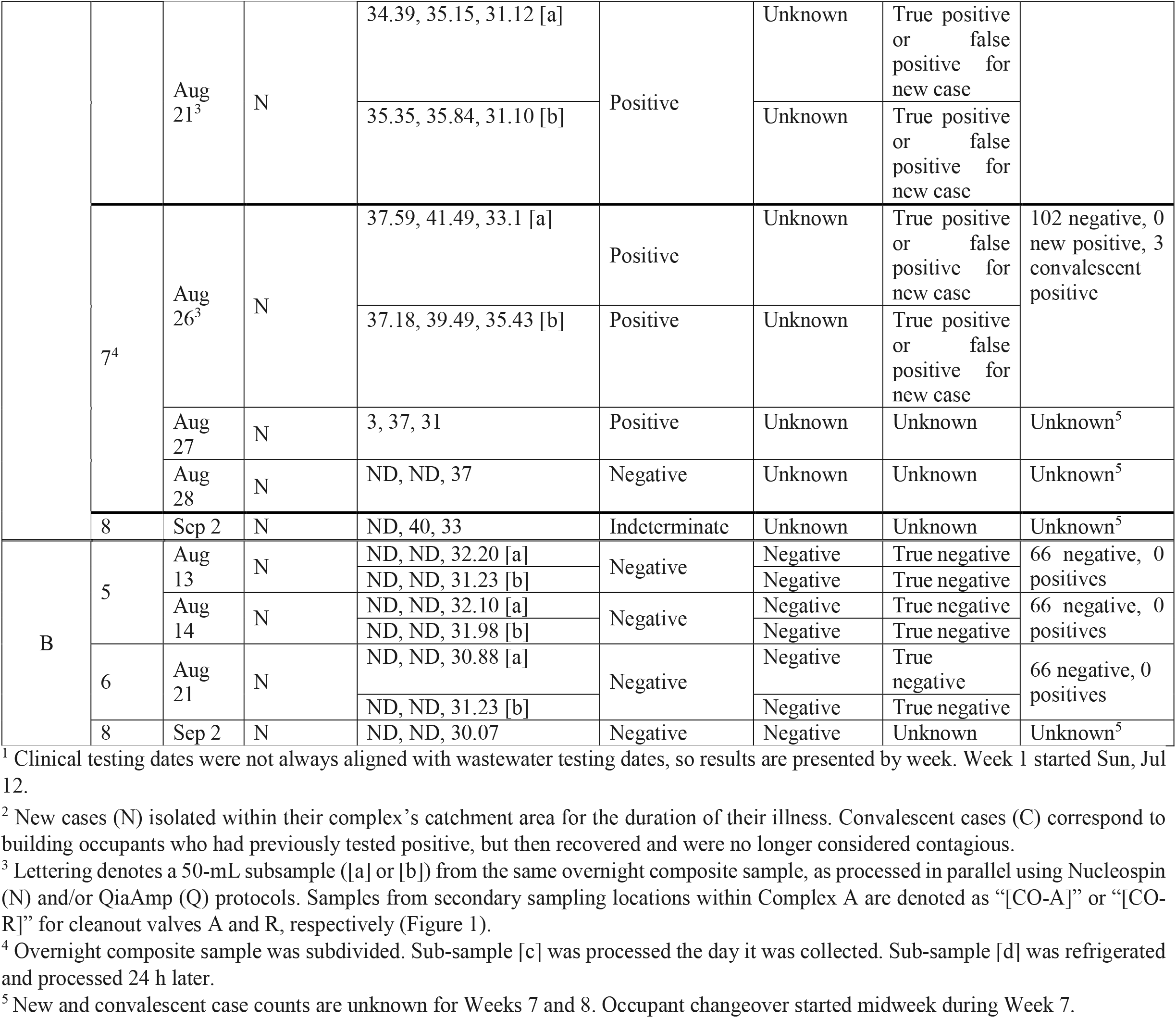
Sampling results for two UVA dorm complexes over time. Lower Ct values indicate higher detected concentrations of SARS-CoV-2. Extraction methods are NucleoSpin (N) and QiaAmp (Q). Not detected (ND), which indicates Ct > 45. Unless otherwise noted, samples were collected from the manhole locations identified in Figure 1. New cases are newly positive in a particular week before the sampling time point and prior positive is thoes who tested positive in weeks prior but still lived in the residence.

**Table 4.** Correlation coefficient (R) values for comparison of Ct values (N1, N2) and corresponding case counts by date. H is “hospital”, RI is “raw influent”, PS is “primary solids”.

| Comparison | R |
| --- | --- |
| N1 vs. N2 - H only | 0.92 |
| N1 vs. N2 - H + RI | 0.94 |
| N1 vs. N2 - H + PS | 0.93 |
| N1 vs. N2 - H + RI + PS | 0.94 |
| N1 vs. Cases - H only | -0.22 |
| N1 vs. Cases - H + RI | 0.31 |
| N1 vs. Cases - H + PS | 0.17 |
| N1 vs. Cases - H + RI + PS | 0.32 |
| N2 vs. Cases - H only | -0.18 |
| N2 vs. Cases - H + RI | 0.41 |
| N2 vs. Cases - H + PS | 0.16 |
| N2 vs. Cases - H + RI + PS | 0.36 |

It was observed that the detectability of SARS-CoV-2 decreases with increasing duration between sample collection and processing. Although the number of samples is small, Ct values for sub-samples of hospital and dorm wastewater processed on the day of sample collection were slightly lower than Ct values for the same samples processed one day later. Relevant samples for both preliminary comparisons, NucleoSpin versus QiaAmp and same-day versus next-day processing, are highlighted in Tables 2 and 3.

Finally, results from additional secondary sampling locations within Complex A (i.e., cleanout valves A and R) reveal that is it possible to monitor multiple locations within a single building at the same time. However, the clean-out valve A was only sampled at a single time point there was a known positive occupant within 14 days of a positive test in building 4 but no SARS-CoV-2 viral targets were detected. Of note during this time period in the clean-out valve the overall flow was very low in volume. Monitoring the building (cleanout value R) with no positive known occupants within the complex gave consistently negative results.

### Validation and Interpretation of Wastewater Results

Ct measurements for the different wastewater sample types were compared with known Covid-19 case counts over time within each sampled building or catchment area. The goal of these comparisons was to: 1) assess the validity of positive/negative determinations arising from wastewater testing; and 2) evaluate whether numerical Ct values and case counts over time are mathematically correlated. During the testing period for the inpatients the average time from symptom onset of and positive PCR was 15 and 11 days respectively and thus there were patients within the contagious period (0-14 days from symptom onset/PCR positivity) ^16, 18^ from all hospital time points and thus all wastewater was anticipated to be positive from infectious cases. There were no new cases in Complex A after week 2 and thus the infectious period of all occupants would have ended by week 4.

If we evaluate only the Nucleospin conclusive results across hospital and dormitory and compare to the cases where the test should have been positive and negative based on the infectious period occupant data (20 true positive, 0 false positive, 1 false negative and 11 true negatives) we determine a sensitivity of 95% and a specificity of 100%. If all the cases of persistent shedding are considered true positives (n=30) then sensitivity increases to 96.9% with 100% specificity by including the additional weeks 4-7. However, if the detection of convalescent shedding is considered a false positive then (n=10) and the sensitivity would be 95.2% but the specificity would be 52%. This highlights the inability of this method to distinguish new contagious infections from persistent viral shedding.

The timing of the transition from positive to negative coincides with the changeover of building occupants, which started midweek during Week 7. There was a lack of clarity at the individual level around movement and thus week 8 was excluded from analysis from both complex A and B. All students were tested immediately upon arrival and instructed not to return to campus if symptomatic. This presumption is consistent with negative wastewater results during Week 8.

With our current method there was no obvious mathematical correlation between documented Covid-19 prevalence and Ct value of the wastewater results. There was very weak negative correlation (R = −0.2) between Ct values and known hospital case counts by date, for both N1 and N2. This correlation did not hold when either or both set of WWTP samples was also considered. Ct values for the N1 and N2 amplicons were highly correlated with each other (r ∼0.94) across each kind of wastewater sample (hospital, raw influent, and primary solids) and for all samples combined.

The small number of positive cases occurring in the sampled dorms during this study precludes rigorous statistical comparison between measured Ct values and corresponding case counts by date. However, it is notable that no new cases were recorded in Complex A after Week 2. Therefore, the positive results obtained during Weeks 3-7 are attributed to persistent viral shedding from a relatively small number of convalescent occupants (3 out of 105). There was no apparent trend in Ct values over time since discovery of the last new case. That is, there was no indication that Ct values gradually increased starting in Week 3 to 7 as known cases convalesced.

## Discussion

### Collecting representative wastewater samples

In summary, we demonstrated an accessible method for performing wastewater surveillance on congregate settings with external validation of the results based on occupant testing. Although this is a small data set there are several lessons which are important to note for both the method and the interpretation. First, it was of interest to know how population size affects SARS-CoV-2 detectability in occupied residential buildings. Practically, UVA Hospital and most UVA dorms offer multiple sampling locations, such that it was possible to monitor different population sizes within the same building. We had an instance where a positive case was missed among a 15 person occupancy. This may have been because of inadequate sample collection due to low water use by less than 20 occupants or simply because of the positive case had a bowel movement which was missed. However, we took this to indicate that our current method is best targeted at building occupancy of 20 or more persons. However, it is likely possible to sample much larger populations. Recently published studies of community-level wastewater-monitoring for SARS-CoV-2 have estimated that the sensitivity of this approach may be as low as 1 positive case per 10,000 ^19^.

From a practical standpoint, dye testing was an important means of confirming that wastewater from a particular building or wing flows to a candidate testing site. This also confirmed the very short residence/travel times for individual toilet flushes. Ultimately we had started with grab sample collection at a single time point (data not shown) but due to the widely varying daily schedules of building occupants, coupled with the short resident time of outflow composite sampling seemed more appropriate for the purpose intercepting all potential viral RNA shedding. However, composite versus grab sample was not directly compared in this study. Twenty-four hour collection (but often closer to 22-23 h in practice) yielded good positivity when known Covid-19 cases were present among building occupants. However, shorter compositing durations may be feasible, depending on contextual factors (i.e., occupant daily schedules, catchment flow rate and travel time to the sampled location, etc.). One 11-h composite sample was collected from the hospital cleanout valve and results were not noticeably different from samples collected using the longer durations from the same site.

Sample handling after collection will likely also be an important factor. In a complex sample such as wastewater, mitigating viral RNA degradation over time is critical to maximizing method sensitivity and SARS-CoV-2 detectability^23, 25^. This study reports on collected during the summer, when daytime temperatures were routinely as high as 38 °C (101 °F), using “portable” autosamplers which do not come with wall outlet power adaptors. Because it was not possible to refrigerate the autosamplers while they were in use, sample jars were packed on ice during sample collection and transport to lab. This approach seems to have been adequate. However, there are now several firms offering fee-based SARS-CoV-2 analysis for clients who ship their wastewater samples to centralized testing facilities. Limited examination of RNA detectability over time (same-day processing versus 1-d later) suggests that long travel times could reduce viral RNA detectability, which could decrease the usefulness of wastewater-based surveillance as an early warning system for SARS-CoV-2 outbreaks in congregate living settings ^40^.

### Comparing and optimizing molecular diagnostic techniques

This study initially focused on sampling from a Covid-19 tower at the UVA hospital and a local municipal WWTP because it was expected that these locations would yield samples with detectable quantities of SARS-CoV-2. This was critical for comparing and refining molecular methods from literature before sampling from occupied buildings.

Effective RNA concentration is critical to the overall success of the proposed approach ^39, 41^. Ultracentrifugation was roughly as effective as the electropositive filter method in this study ^27^, which was somewhat unexpected based on existing literature ^41^. However, this could be because electropositive filtration targets negatively charged viral particles, whereas this approach makes use of RNA fragments rather than intact virus. ^32, 42^. Because the performance of these two methods was not dramatically different, the decision to move forward with ultra-centrifugation came down to practical consideration of its benefits and drawbacks. Regarding benefits: the method is highly sensitive, consumes very small sample volumes, and does not require specialized reagents or costly consumables that may be intermittently difficult to procure during the pandemic. A critical drawback is that ultra-centrifuges are very costly. Also, because centrifugation is a batch process (i.e., only a fixed number of samples can be spun at the same time), and long spin-up and spin-down times are required to achieve the high g-forces needed to settle viral particles, this approach is only moderately scalable. As a result, individual labs may struggle to process the large numbers of samples arising from testing multiple buildings multiple times per week. As we have little head-to-head data the major drawbacks to the elecropositive filtration methods were practical; the supply chain issues on obtaining the filters as well as cost, personnel time biohazard risk and logistics of filtering 6 L of wastewater. The other methods of RNA concentration with filtration and precipitation were not as effective and thus not further pursued however, it should be noted that the components of these protocols and methods which could have been refined for improved recovery were not further explored and that the RNA extraction method seemed very impactful and could have explained some of the differences ^19, 28^.

Effective RNA extraction is also a critical step in managing a complex specimen such as wastewater given how many potential PCR inhibitors are present in wastewater ^23, 28, 42^. We however, only thoroughly compared two commercial kits. It was observed that NucleoSpin worked better than QiaAmp for the samples collected in this study with less inhibition but it should be noted that there were still inhibited samples. In addition, without a quantified extraction control it is not possible to evaluate the degree of partial inhibition from any given sample ^43^. Finally, although the results are not presented in detail, samples from this study were run on multiple commercial PCR platforms, such as Abbott Allinity and m2000. However, it was observed that samples routinely failed on commercial platforms with “built-in” extraction protocols. These failures were attributed to inhibition caused by wastewater constituents ^23^. This observation highlights the need for careful evaluation of RNA extraction protocols as key to ensuring successful amplification and good overall method sensitivity.

Wastewater samples collected in this study were run together with clinical samples from UVA hospital (and elsewhere), using the same PCR platform. This platform is based on the CDC protocol for SARS-CoV-2 analysis, which was not originally intended for wastewater-based testing^34^. However, it was convenient for UVA to process its clinical and wastewater samples together, subject to the same quality control procedures, because testing equipment and trained personnel were already in place. Clinical diagnostics require very high reliability and sensitivity, which are also extremely valuable in the context of pooled wastewater-based surveillance for congregate living facilities (e.g., to get ahead of an outbreak by finding a small number of positives within the overall building population).

### Validating and interpreting results

Ultimately this most important outcome of this study was in having a known group of asymptomatic students in a congregate setting and symptomatic hospitalized patients and demonstrate that the method was sensitive and aligned with the known carriage of the building occupants Covid-19 infection status when there is enough wastewater flow. The method was also reproducibly positive on a community with known cases, thereby validating a reasonable limit of detection for a larger population.

The goal of this surveillance method would ultimately be paired with testing all individuals in a building once a new contagious positive is detected. The method used in this study gives reliably positive results even when the number of positive (or convalescent) cases in a sampled population is very small. However, the downside of using this very sensitive molecular detection method was that it could not distinguish new cases from persistent convalescent shedding. It is well documented that persons shed SARS-CoV-2 RNA for weeks in their stool after infection ^17, 44^. This is evident from the Ct measurements over time at Complex A, whereby viral shedding from a very small number of asymptomatic cases was detectable for several weeks after the last new case was identified. Results stayed positive (Ct < 45) for more than a month, with no discernable indication that viral quantity was decreasing over time. We hypothesized that the Ct detection would increase as post-infectious shedding resolved however with our non-quantitative method we did not see an indication of this in the data. In addition, the observed Ct values cannot be used to impute what number of newly positive cases from convalescent shedding are present in a particular building at any given time. It is anticipated that the SARS-CoV-2 RNA stool shedding would decrease overtime and thus be reflected in higher Ct values with no new cases^16, 17^. Possible strategies for overcoming this challenge may include refinement of the molecular method to make them more robustly quantitative or application of statistical modeling approaches, once there is better data to parameterize fecal viral shedding over time for symptomatic and asymptomatic cases.

In conclusion, results from this pilot study confirm that we have developed a feasible method for wastewater-based testing at individual building level as a means of implementing passive pooled SARS-CoV-2 surveillance for occupied congregate living settings. Because we are attempting to use this as a method of pooled surveillance on congregate settings we wanted sensitivity over accurate quantification. The molecular methods used in this study, based on literature and existing clinical methodology, are highly sensitive but not specific for new infections. Persistent convalescent shedding will likely be an important technical challenge that must be overcome in future work so that the proposed approach can be successfully deployed in congregate living settings such as skilled nursing homes, university dormitories and prisons.

## Data Availability

All data available and is presented in the article without other data repository.

## Acknowledgements

This work was funded by a UVA Engineering in Medicine Seed Grant, and support from the University Reopening Committee. The authors also express their sincere gratitude to the following individuals: Professors Teresa Culver and James Smith for use of their autosamplers; Mr. Robert Haacke, Mr. David Tungate, and Ms. Jennifer Whitaker of RWSA for their technical advice and assistance in collecting WWTP samples. Thank you to Roland Zumbrunn for technical and logistics assistance on student movement and occupancy rates.

